# Life Expectancy Exploration of Heart Failure Patients Presenting with Diabetes

**DOI:** 10.1101/2024.09.03.24312945

**Authors:** Prosper Narh, Michael Asante Ofosu, Elliot Owusu Addo, Daniel Wiafe Preko, Grace Aba Bart-Plange, Roselyn Oforiwaa Acquah

## Abstract

**Background:** Diabetes is a serious medical condition marked by high blood sugar levels, leading to complications in organs like the kidney, liver, and heart. Previous research has identified diabetes as a primary risk factor for congestive heart failure (CHF), a condition where stiffened heart muscles hinder oxygenated blood circulation. Despite its severity, few studies have examined CHF prognosis in diabetic patients. This study aims to provide survival estimates and provide their comparisons among predictors, assess mortality risks based on specific variables, and reveal patient outcome patterns.

**Methods:** We analyzed data from a 2015 study at Faisalabad Institute of Cardiology and Allied Hospital (FICAH), Pakistan, focusing on 125 diabetic patients with Class III or IV heart failure and left ventricular systolic dysfunction. Kaplan-Meier estimates provided survival rates, log-rank tests compared survival across variables, Cox regression models estimated hazard ratios, and cluster analysis grouped patients by characteristics and survival rates.

**Results:** The study examined 125 diabetic heart failure patients. Of these, 40(32%) were censored and the remaining 85(68%) died. The total follow-up period was 278 days, on the 120^th^ day, 0.752[95% CI = 0.678,0,883] of the patients survived after initial diagnosis. Log-rank tests showed survival differences linked to high blood pressure, anemia and smoking. Cox regression identified age, smoking and hypertension as major mortality risk factors. Cluster analysis revealed the risk factors associated with middle aged and the older patients.

**Conclusion:** The overall prognosis of heart failure patients with diabetes was poor with high mortality rates, implying effective treatment and management.

## INTRODUCTION

Diabetes and heart failure are two interconnected conditions that have a profound impact on each other’s progression and outcome. The coexistence of diabetes and heart failure creates a vicious cycle where each condition worsens each other, diabetes damages the heart and blood vessels increasing heart failure, while heart failure exacerbate diabetes by promoting insulin resistance and hyperglycemia [1]. In patients with congestive heart failure (CHF), diabetes is associated with a worse prognosis including mortality, and the presence of diabetes in CHF patients is linked with a higher risk of cardiovascular events such as stroke [2].

Heart failure is a serious health care problem not only for patients and their family but also for society, as it contributes significantly to the enormous costs associated with the care of affected individuals. Nearly 6.5 million people in Europe, 5 million people in the United States (US), and 2.4 million people in Japan suffer from heart failure [3]. Also, diabetes is a serious and costly public health problem in the US affecting more than 16 million people [4]. Previous research conducted by Kennel et al. pinpointed diabetes as the major cause of congestive heart failure (CHF) [5], as such diabetic heart failure becomes a major concern for public health and individuals with this condition. The prognosis of heart failure is poor with reported survival estimates of 50% and 10% at 5 and 10 years respectively [6]. Also, approximately 75% of diabetics die from vascular complications leading to heart failure [7].

The cost of medical treatment for diabetes and heart failure is alarming in our world today, due to this, more investigation is done concerning this medical condition. For instance, in Ireland, the average cost of hospital admission for heart failure is IR 2146 pounds [8] and diabetes cost in the United States estimated $132 billion in 2002 as a result of medical costs and lost productivity [9].

Although there have been a lot of studies on diabetes and heart failure, few studies have focused on how diabetes affect the survival in HF patients, therefore this study seeks to understand the prognosis of heart failure patients with diabetes. Specifically, we aim to (a. provide the survival estimates and provide comparisons among certain patients’ characteristics, (b. determine the relationship between some patient characteristics and the risk of death from HF among diabetic patients and (c. identify patterns of patient outcomes based on their survival rates and characteristics.

## MATERIALS AND METHODS

### Data source and study design

The data used in this research was secondary and was obtained from previous research conducted by Davide et. al [10]. Data from their research were extracted from the Faisalabad Institute of Cardiology and at the Allied Hospital in Faisalabad (Punjab, Pakistan) from April 2015 to December 2015, their study contained 299 patients with congestive heart failure (CHF). They aimed at applying several machine learning classification models to predict the survival of heart failure patients, and also rank the features corresponding to the most important risk factors of heart failure patients. Their study’s dataset included demographic and medical records of 299 heart failure patients.

The dataset included information on time until death making it suitable to address our research’s objectives. In this study, we specifically concentrated on heart failure patients with diabetes. Non-diabetic patients were excluded from the dataset making the final sample size of 125 patients. Our study was based on the fact that diabetes and heart failure are interconnected medical conditions and the condition of one worsens the other. As such patients presenting with both conditions need to know the risk associated and seek medical advice.

### Variables

In our analysis, we employed the dataset from Faisalabad Institute of Cardiology and at the Allied Hospital in Faisalabad (Punjab, Pakistan). Within the dataset, we made use of the ‘time’ variable which represented the patients follow up time and ‘status’ which determined whether the patient is censored (survived) or not censored (died) at the end of the study. Follow-up time used in this study is the duration (days) in which the patient is tracked until the event of interest (death) occurs.

Additionally, our dataset involved demographic and health related information, including the patients age, sex, smoking status, anemic status, high blood pressure, ejection fraction, creatinine phosphokinase, sodium creatinine, serum sodium and platelet levels. Smoking status, anemia and high blood pressure were grouped as categorical variables where “0” represented “absent” and “1” represented “present”. Also, sex was grouped as ‘0’ and ‘1’ representing women and men respectively. Numerical variables were time, creatinine phosphokinase, ejection fraction, serum sodium, serum creatinine and platelet levels.

### Statistical analysis

Descriptives like frequencies and ratios were used to illustrate describe categorical data. Quantitative data were described using the mean, median and mode. Survival times were calculated from when patients first joined the study. Kaplan-Meier estimates were used to calculate survival probabilities, and Log-rank tests were used to compare survival rates across different groups.

The Cox Proportional Hazard (PH) model was utilized to estimate hazard ratios and determine the relationship between patient characteristics and the risk of dying from HF as a diabetic patient. A hazard ratio over 1 indicates increased odds for a specific category. Percentage interpretation involves computing the percentage change in hazard for every one-unit predictor change. For instance, a hazard ratio of 1.60 implies a 60% rise in risk, while 0.30 indicates a 70% decrease, holding other factors constant. Cluster analysis examined survival patterns among patients based on their survival durations and predictor variables like smoking status, anemia, age and high blood pressure. A significance level of 0.05 was maintained throughout the study. Statistical analyses were conducted using R statistical software (version 4.3.2).

## RESULTS

### Descriptives

Our study population consisted 125 diabetic heart failure patients. Out of the 125 patients, 70 of them were males and the remaining 55 of them females (Table 1). 68% (85) of the patients died from heart failure and the remaining 32% (40) survived(censored) (Table 1). The minimum and maximum follow-up times were 0 and 278 days (9.3 months) respectively (Table 2). The minimum and maximum ages were 40 and 94 years respectively (Table 2) highlighting the high prevalence of heart failure in middle aged and older adults.

**Table 1.** Frequency distribution of participant characteristics.

| Variables | Frequency | Percentage |
| --- | --- | --- |
| Sex: 1 | 70 | 56 |
| 0 | 55 | 44 |
| Anemia: 1 | 53 | 42.2 |
| 0 | 72 | 57.6 |
| HBP: 1 | 43 | 34.4 |
| 0 | 82 | 65.6 |
| Smokers: 1 | 30 | 24 |
| 0 | 95 | 76 |
| Death: 1 | 85 | 32 |
| 0 | 40 | 68 |

*HBP – High Blood Pressure.*
*Right censoring is assumed in this study.*
*For Sex, “1” represented males and “0” represented females, for anemia and HBP, “1” represented disease present and “0” represented disease absent, for smokers and death, “1” represented event happened and “0” represented event did not happen.*

**Table 2.** Summary statistics of participant.

| **Variables** | **Mean** | **Median** | **Minimum** | **Maximum** |
| --- | --- | --- | --- | --- |
| Age (Years) | 59.42 | 60.00 | 40.00 | 94.00 |
| Time to-follow-up (Days) | 133.3 | 120.0 | 0.0 | 278.0 |

Comorbidity profiles included anemia and high blood pressure. For anemic status, non-anemic cohorts comprised of 72 (57.6%) individuals while anemic cohorts comprised of 53 (42.4%) individuals. This emphasizes that non-anemic cohorts have better clinical outcomes like fewer hospitalization and improved functional capacity than anemic patients. Also, non-high blood pressure cohorts consisted 82 (65%) patients while high blood pressure cohorts included 43 (34.4%) patients (Table 1). This signifies that greater proportion of patients have a well-controlled blood pressure and healthier life style.

### Survival estimates and log rank tests

Survival probabilities of diabetic heart failure patients were estimated at different points using Kaplan-Meier estimates (Figure 1). On the eighth day of the follow-up, the survival probability was 0.992(95% CI= [0.977,1.000]), meaning that approximately 99% of patients survived after day 8. Exactly after 120 days (four months) of follow-up, the survival probability of patients was 0.752(95% CI= [0.678,0.883]), which signifies that about 75% of diabetic heart failure patients would survive past the first fourth months after initial diagnosis. The survival probability continues to decrease slowly until the end of the study (278 days (9.3months)) where about 40% of the patients survived (Table 3).

**Figure 1.**
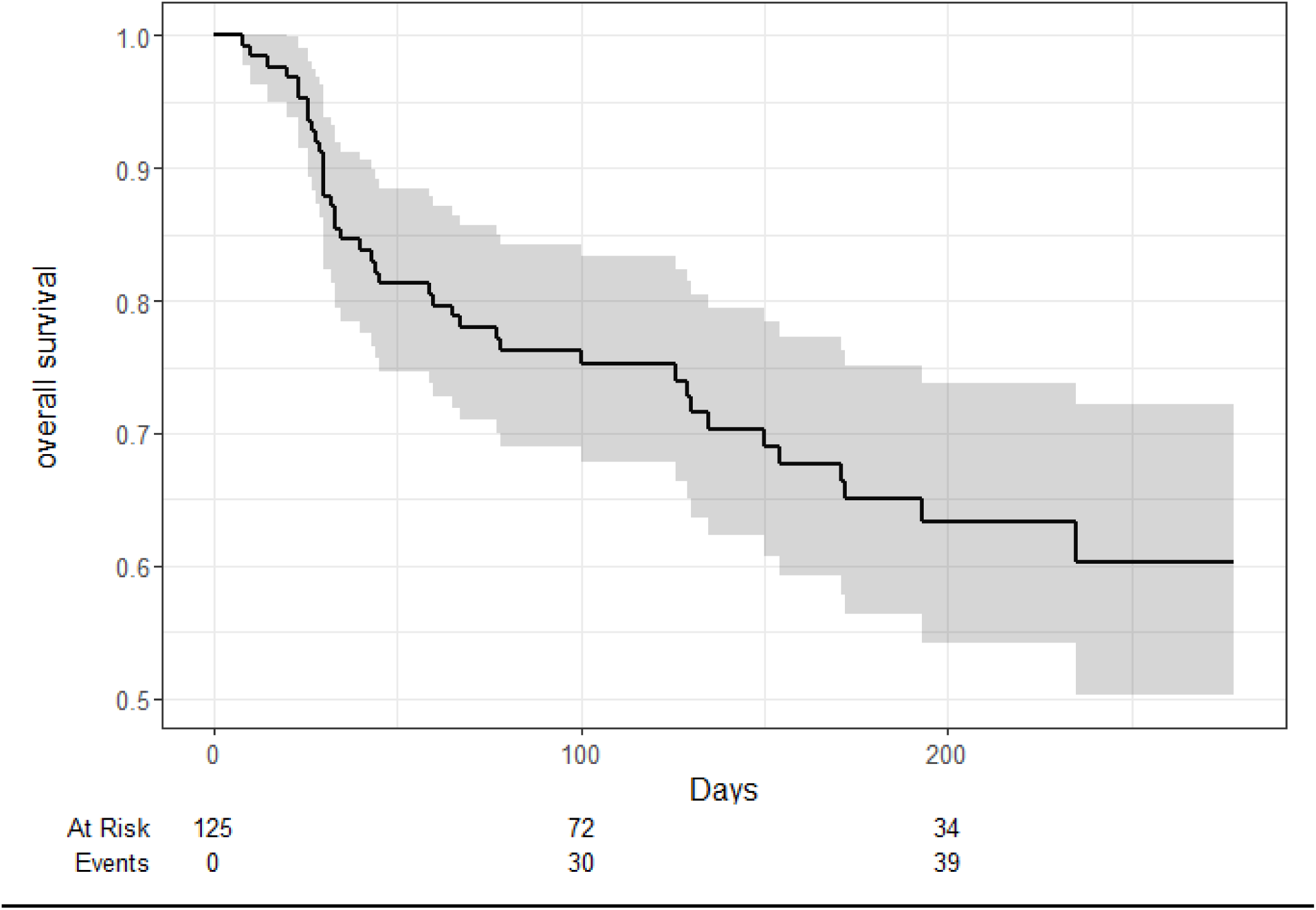
Kaplan-Meier curve for diabetic heart failure patients

**Table 3.**
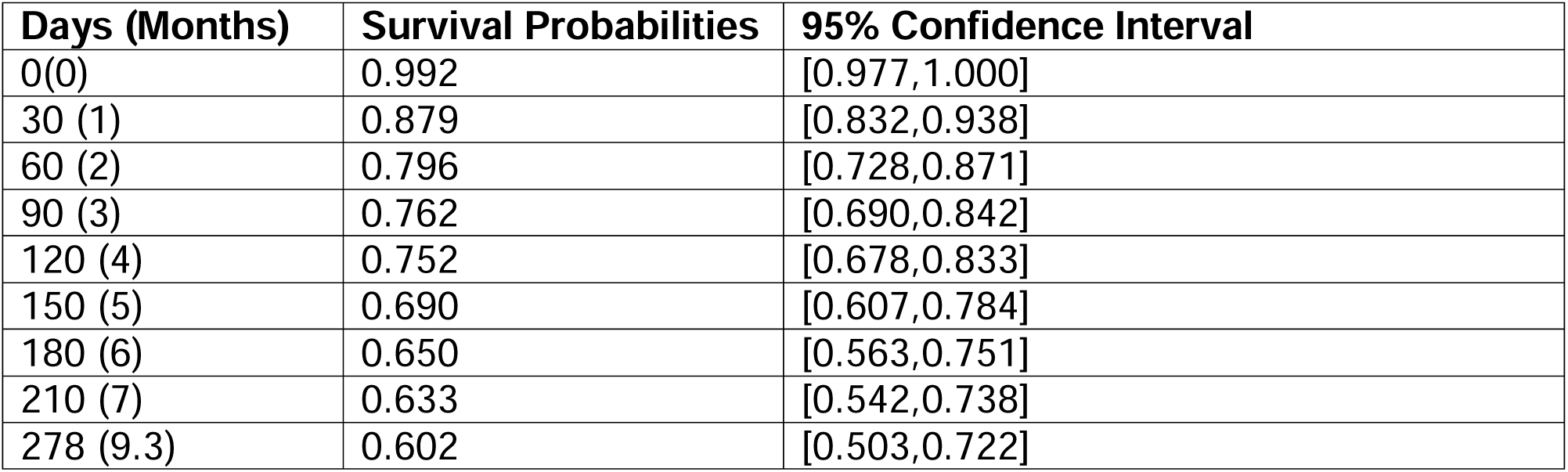
Survival estimates for patients.

In Table 4, the log rank test was used to compare categorical variables based on their survival probabilities. For smokers and non-smokers, the p-value from the log rank test was 0.002 which suggested that there was a statistically significant difference between the survival of patient who smoked versus those who did not smoke. This signifies the fact that smokers who are diabetic heart failure patients die faster as compared to the non-smokers. Based on sex(gender), the p-value from our log rank test was 0.04. This implies that there is a statistically significant difference between the survival rate of both sexes, highlighting that males and females have a different rate of surviving heart failure in diabetic patients. Also, for high blood pressure and non-high blood pressure cohorts, 0.01 was the p-value obtained from the log-rank test, which again means there is a statistically significance difference between the survival rates of these two groups. This supports the fact that high blood pressure is a prognostic factor for poor outcomes in patients presenting with diabetes and heart failure, unlike non-high blood pressure patients. Conversely, for anemic and non-anemic cohorts, the p-value obtained was 0.6 which signified that there is no statistically significance difference between those with anemia and those without anemia. This may be because anemia is not a significant predictor of diabetes progression hence the indifference in the survival rates.

**Table 4.** Summary of log-rank test.

| Variables | Log-rank statistic | Chi-square statistic | P-value |
| --- | --- | --- | --- |
| HBP (Absent)<br>(Present) | 0.808<br>1.827 | 2.67<br>2.67 | 0.001 |
| Anemia (Absent)<br>(Present) | 0.0967<br>0.1378 | 0.236<br>0.236 | 0.6 |
| Smoking (Absent)<br>(Present) | 0.296<br>1.025 | 1.33<br>1.33 | 0.002 |
| Sex (Female)<br>(Male) | 0.126<br>1.129 | 0.70<br>0.70 | 0.04 |

### Cox-proportionnal hazard model

In the Cox PH model for diabetic heart failure patients in Figure 2 and Table 5, smokers had a significantly higher risk of death (HR = 2.228) compared to those who do not smoke, and the p-value associated with this risk was 0.0669. This means that the hazard rate of 2.228 suggest a 122.8% increase in the risk of death for smokers compared to non-smokers and the p-value of 0.0669 indicate that the observed difference in hazard rate between these two groups is not statistically significant. High blood pressure (HBP) also showed a relatively high risk of death (HR = 1.663) and p-value of 0.1639. The hazard rate implies that HBP is a major risk factor for congestive heart failure whiles the p-value suggest that HBP is not statistically significant. However, sex(female) had a lower risk of death on diabetic heart failure patients (HR = 0.4022) and a p-value of 0.0272. This signifies that there is about 60% decrease in the risk of death for females. Hence the male sex is rather at risk of dying from heart failure. Creatinine phosphokinase and blood platelets had no effect on death as their hazard rate was 1, holding all other predictor variables constant.

**Figure 2.**
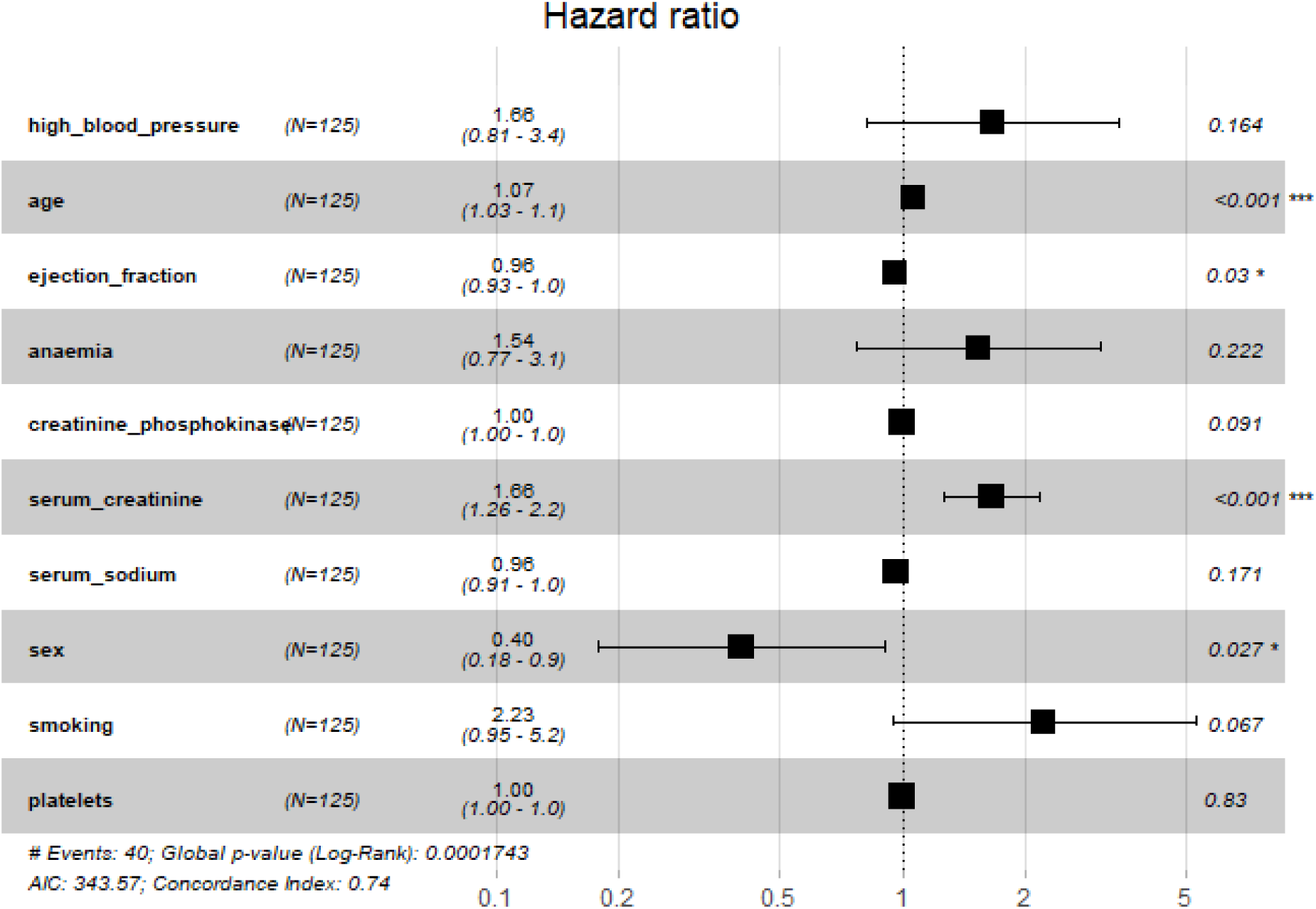
Hazard ratio of patients

**Table 5.** Cox-Proportional Hazard Model.

| Variable | Coefficient | Hazard Ratio | P-value |
| --- | --- | --- | --- |
| HBP (Present) | 0.5089 | 1.663 | 0.1639 |
| Age | 0.06336 | 1.065 | 0.0006 |
| Ejection Fraction | -0.03891 | 0.9618 | 0.0305 |
| Anemia (Present) | 0.4322 | 1.541 | 0.2219 |
| CP | 0.000306 | 1 | 0.0905 |
| SC | 0.5047 | 1.656 | 0.000312 |
| Serum Sodium | -0.03751 | 0.9632 | 0.1714 |
| Sex (Females) | -0.9109 | 0.4022 | 0.0272 |
| Smoking (Smokers) | 0.8010 | 2.228 | 0.0669 |
| Platelets | -0.0000004 | 1 | 0.8303 |

*HBP – High Blood Pressure; CP – Creatinine Phosphokinase; SC – Sodium Creatinine*

### Cluster analysis

The presence of two distinct clusters (Figure 3) indicates that the data points are separating into two distinct groups or populations based on the two dimensions (Dim1 and Dim2) plotted. The red cluster (cluster 1) is significantly larger than the teal cluster (cluster 2). This implies that the majority of data points belong to the population represented by cluster 1, while cluster 2 is smaller, potentially an outlier group. The teal cluster 2 is tightly concentrated and the red cluster 1 is more dispersed, indicating greater variability within this population, especially along the Dim1 axis.

**Figure 3.**
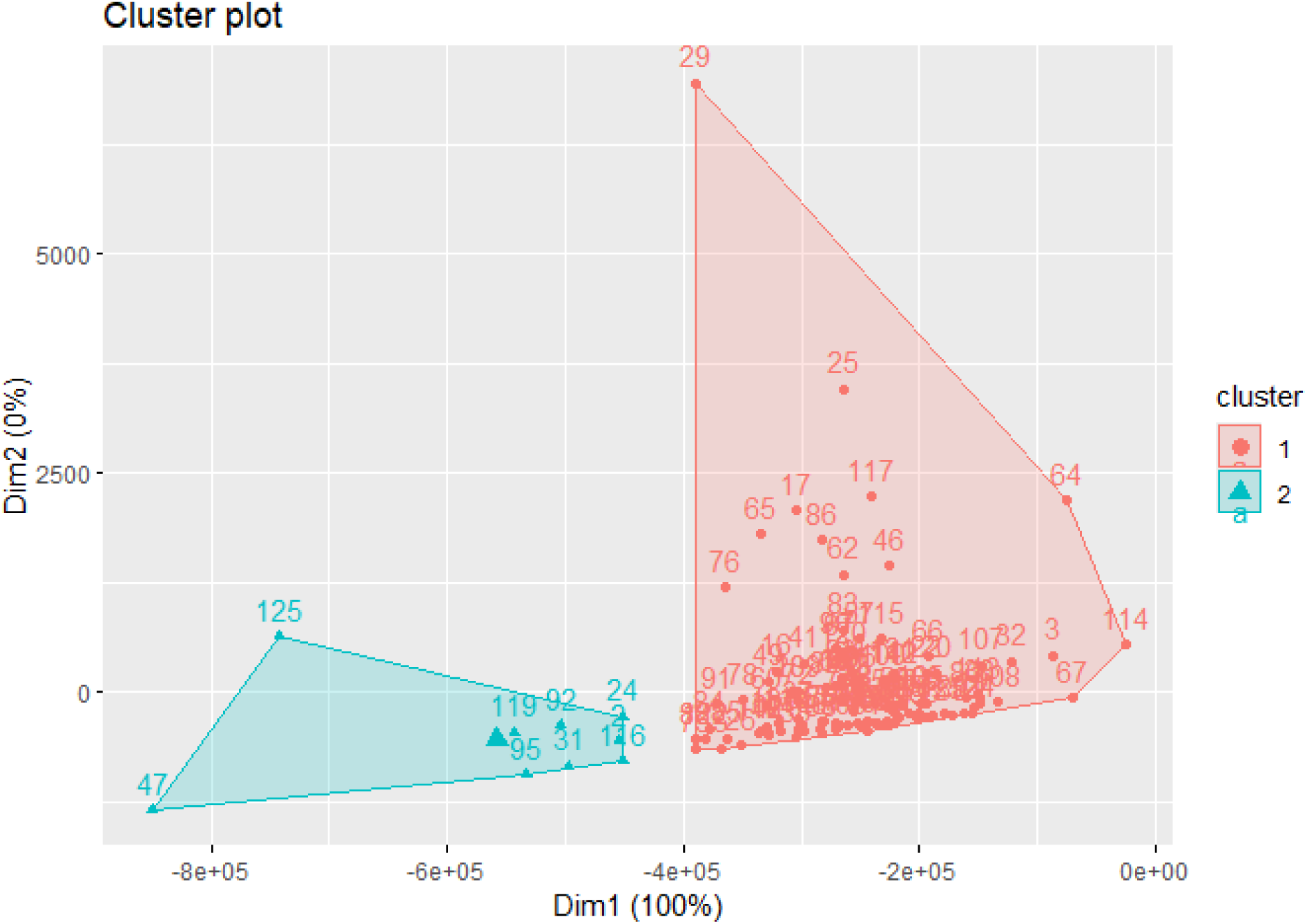
Cluster plot based on classification of survival rate

Cluster 1, comprising predominantly older individuals with a higher prevalence of hypertension (HBP), anemia, and elevated levels of creatinine phosphokinase. This reflects a subgroup potentially burdened by multiple comorbidities and greater cardiovascular risk factors. Cluster 2 on the other hand, exhibits a younger age profile with a higher representation of males and smoking habits, suggesting a distinct demographic makeup.

## DISCUSSIONS

The main findings from this study were (a) there is a statistically significant difference in the survival of smokers and non-smokers in diabetic heart failure patients likewise high blood pressure and non-high blood pressure cohorts. This signifies that smoking and high blood pressure constitute as a major risk factor in the survival of this patients. (b) Age, high blood pressure and smoking had a high rate of death on diabetic heart failure patients while sex had a lower rate of death.

In our study, we found that smoking and high blood pressure were associated with a relatively higher hazard rates compared to the other predictor variables, this aligns with previous researches. Hypertension has been identified as a major risk factor for heart failure and a common comorbidity in diabetes [12]. The combination of high blood pressure and diabetes increases the risk of damage to the heart resulting in heart failure [13,14]. Haire et al. [11] and Kathleen et al. [15] found smoking as a primary risk factor for heart diseases and type II diabetes in their respective studies. This aligns greatly with our study’s findings, where smoking was significantly associated with increased risk of death from HF, further indicating smoking’s bad effect on the body’s organs. Kalogeropoulos et al. [16] and Nunez et al. [17] found that creatinine phosphokinase and platelets levels have no effect on mortality of patients with heart failure. This confirms our findings where both clinical indicators had a hazard rate of 1.

Smokers and non-smokers in our study had different survival rates on heart failure and diabetes. From our log rank test, non-smokers had a better rate of survival compared to smokers. A study by the World Health Federation confirms that, smokers have a significantly lowers rate of survival as compared to non-smokers and the risk of heart failure is 2-4 times higher in smokers than in non-smokers [18]. High blood pressure cohorts and non-high blood pressure cohorts also had a significantly different survival rates, with non-high blood pressure cohorts having the better survival of patients presenting with heart failure and diabetes. A study by Tsimploulis et al. (2017) found that non-hypertensive heart failure patients exhibited a 5-year survival rate of 75%, whereas hypertensive patients had a markedly lower survival rate of 60% [19]. Hypertensive patients are often burdened with comorbidities such as diabetes and chronic kidney disease, which further complicate heart failure management and adversely affect prognosis [20]. For sex, we found that males and females have unequal survival rate of heart failure and diabetes. A recent study by Adams et al., found that there is a gender difference in the survival rate of heart failure, where the female gender lives much longer compared to males and suggesting that men and women have incomparable prognoses [21].

In our cluster analysis, we found that older patients often have high blood pressure and anemia in cluster 1. A study by Thomas et al. [22] and Alpert et al. [23] stated that hypertension and anemia are found in patients who present to the hospital with diabetes and advanced heart failure. This signifies that in cluster 1, older patients have greater risk of dying faster since both HBP and anemia have a very high hazard rate from our cox-proportional model. In cluster 2, we found that middle aged adults have low high blood pressure and frequent habit of smoking. Studies by Rodgers et al. [24] and Joes et al. [25] confirm this, noting that the young and middle-aged adults usually have fewer health issues, better heart function, and more physical resilience to cope with heart failure.

### Limitations

Our research study had several limitations that must be acknowledged. First, the dataset we analyzed contained a relatively small number of patients who had both diabetes and heart failure. This may be because it was difficult to obtain data on patients with both medical conditions. Having a smaller sample size limits how much we can generalize or apply our findings to broader populations.

Secondly, our dataset grouped together all heart failure patients who also had diabetes. We did not specify whether the patients had type 1 or type 2 diabetes. Separating and analyzing the two diabetes types may have provided more useful information, since we know there are differences in the risk factors associated with each type.

Lastly, because we used an existing dataset collected by others, we had to make some modifications to the original data. This data alteration process may have inadvertently changed or introduced errors into a key variable we used for analysis. As a result, there could be some confounding variables we did not properly account for that may have impacted our results.

These limitations mean the results from our study should be interpreted with some caution. Further investigation through additional research may be needed to validate and expand on our findings, especially with a larger sample size that distinguishes diabetes types. Despite the limitations, this study still provides insights into factors impacting outcomes for patients with both diabetes and heart failure.

## CONCLUSION

In summary, this study provides valuable insights into the factors influencing survival outcomes for patients suffering from both diabetes and heart failure. From our findings, we identified some key determinants that can guide clinical management and risk stratification. Our study reveals significant differences in survival probabilities among some predictor variables in patients suffering from diabetic heart failure. Modifiable risk factors like smoking and hypertension emerged as significant predictors of mortality, highlighting the crucial importance of aggressive risk factor modification through lifestyle interventions and optimized medical therapy. Females had a better chance of survival and males had the greater risk of dying from diabetic heart failure. Also, from our findings, we revealed that there are two groups of patients presenting with diabetic heart failure, older and middle-aged adults. Older individuals being at risk of dying early from diabetic heart failure and middle-aged adults relatively surviving for a longer period of time. This signifies that ageing is also a risk factor for diabetic heart failure patients. These conclusions carry important implications for clinical practices and management of patient’s specific characteristics. Notwithstanding the limitations of this study, this research warrants more investigations to shed more light on this medical condition.

## Availability of dataset

The data that supports the findings of this research are available under the Creative Common Attribution 4.0 International License (CC BY 4.0). The dataset is accessible through Kaggle database, which provides unrestricted access and reuse of the data. Researchers are encouraged to explore and utilize the dataset while adhering to the terms of the creative common license.

## List of abbreviations

CHF: Congestive Heart Failure

HF: Heart Failure

CI: Confidence Interval

HR: Hazard Ratio

## Data Availability

All data produced in the present work are contained in the manuscript

https://www.kaggle.com

